# LabQAR: A Manually Curated Dataset for Question Answering on Laboratory Test Reference Ranges and Interpretation

**DOI:** 10.1101/2025.06.03.25328882

**Authors:** Balu Bhasuran, Qiao Jin, Angelique Deville, Yonghui Wu, Karim Hanna, Zhiyong Lu, Zhe He

## Abstract

Laboratory tests are crucial for diagnosing and managing health conditions, providing essential reference ranges for result interpretation. The diversity of lab tests, influenced by variables like the specimen type (e.g., blood, urine), gender, age-specific, and other influencing factors such as pregnancy, makes automated interpretation challenging. Automated clinical decision support systems attempting to interpret these values must account for such nuances to avoid misdiagnoses or incorrect clinical decisions. In this regard, we present **LabQAR** (**Lab**oratory **Q**uestion **A**nswering with **R**eference Ranges), a manually curated dataset comprising 550 lab test reference ranges derived from authoritative medical sources, encompassing 363 unique lab tests and including multiple-choice questions with annotations on reference ranges, specimen types, and other factors impacting interpretation. We also assess the performance of several large language models (LLMs), including LLaMA 3.1, GatorTronGPT, GPT-3.5, GPT-4, and GPT-4o, in predicting reference ranges and classifying results as normal, low, or high. The findings indicate that GPT-4o outperforms other models, showcasing the potential of LLMs in clinical decision support.

## Background & Summary

Question Answering (QA) has become a powerful tool in domains like search engines and conversational agents. In the biomedical domain, QA supports efficient knowledge retrieval and management, addressing the needs of both professionals and the general public^1^. Clinicians rely on up-to-date evidence to inform diagnosis and treatment decisions under the principles of evidence-based medicine. Meanwhile, patients increasingly seek to understand their health through accessible, reliable online information^2^. Biomedical/clinical QA systems bridge this gap by enabling structured access to complex medical knowledge.

Automatic QA is a core task in natural language processing (NLP) that involves generating accurate answers to queries based on textual or structured data sources^3^. In the biomedical domain, QA systems are designed to address clinical, research, and consumer health questions using data from scientific literature, electronic medical records, or curated health resources^1^. Over the past decade, several domain-specific QA datasets have emerged, driving advances in machine reading comprehension, information retrieval, and clinical decision support. For example, the BioASQ challenge provides expert-annotated QA benchmarks to evaluate biomedical knowledge retrieval and complex reasoning^4,5^. The PubMedQA^6^ dataset consists of yes/no/maybe questions derived from scientific abstracts, while BioMRC^7^ and CliCR^8^ introduce cloze-style reading comprehension tasks using biomedical articles and clinical case reports. In the clinical setting, emrQA^9^ transforms EHR annotations into over one million QA pairs, facilitating question answering from structured clinical data. Additionally, datasets such as MedMCQA^10^ and HEAD-QA^11^ evaluate multi-subject reasoning using medical licensing-style exams, while SentiMedQAer^12^ and CHQ-SocioEmo^13^ incorporate sentiment and patient-centered perspectives to support emotionally intelligent and socially aware QA systems.

The emergence of transformer^14^ architectures such as Bidirectional Encoder Representations from Transformers (BERT)^15^ and large language models (LLMs)^16^ has renewed interest in applying AI to medical question-answering. Early efforts primarily involved training smaller, domain-specific models such as BioBERT^17^, PubMedBERT^18^, and ClinicalBERT^19^. These models demonstrated steady performance gains on benchmark datasets like MedQA (based on the United States Medical Licensing Examination), MedMCQA^10^, and PubMedQA^6^. More recently, the development of large-scale generative models—such as GPT-4.5^20^, LLaMA 4^21^, Gemini 2.0 Pro^22^, and PaLM^23^—along with their medically adapted versions (e.g., MedPaLM^24^, MedPaLM 2^25^, MedGemma^26^), has driven rapid advancements in medical QA performance. Trained on internet-scale data using massive computational resources, these models have achieved state-of-the-art results on benchmark datasets: Notably, the smaller, medically fine-tuned MedGemma 27B model reached up to 89.8% and MedPaLM 2 up to 86.5% accuracy on the MedQA dataset, while scoring 76.8% and 74.1%, respectively, on PubMedQA. Encouraged by these breakthroughs, researchers have begun applying these models to a variety of clinical tasks, including discharge summary generation^27^, clinical note summarization^28^, clinical trial matching^29^, and differential diagnosis prediction^30^. A critical component underpinning many of these applications is the interpretation of laboratory test results.

Laboratory tests play a pivotal role in healthcare, guiding diagnosis, treatment planning, and monitoring of disease progression^31^. Recently, LLMs such as ChatGPT^32^ have shown promising capabilities in interpreting laboratory test results, a key component of clinical decision-making. Studies by He et al.^33^ and Girton et al.^34^ revealed that GPT-4 and ChatGPT, respectively, provided responses to lab-related questions that were often more accurate and helpful than peer users or even physicians, though limitations in contextual personalization and domain specificity were noted. Meyer et al. expanded the comparison across multiple LLMs (ChatGPT, Gemini, Le Chat) and found that while models achieved near-physician-level accuracy, they sometimes produced overgeneralized or clinically implausible explanations^35^. These findings illustrate the potential of LLMs in lab test interpretation, while also highlighting the need for reliable datasets to support training and comprehensive evaluation. Recent studies have further explored the utility of LLMs in lab test-driven differential diagnosis. Bhasuran et al. demonstrated that incorporating lab data into clinical vignettes significantly improved diagnostic accuracy, with GPT-4 reaching up to 80% accuracy in top-10 differential diagnosis predictions^30^. Similarly, Berg et al. found that ChatGPT’s diagnostic accuracy improved when lab data were included, surpassing physicians in top-5 accuracy for emergency department cases^36^. These results emphasize the clinical value of structured lab data and the importance of domain-specific QA resources. Evaluating the clinical understanding of LLMs—especially their interpretation of lab test results and the reasoning behind clinical outcome predictions—is critically important and remains a highly challenging task. Accurate interpretation of lab results is critical and depends on a range of contextual factors such as age, gender, biological specimen type, and patient-specific health conditions. However, automating this process poses substantial challenges due to the complexity and variability inherent in laboratory diagnostics. To this end, we introduce LabQAR (**Lab**oratory **Q**uestion **A**nswering with **R**eference Ranges), a manually curated question-answering dataset focused on laboratory test reference ranges and interpretation.

LabQAR is designed to capture the context sensitivity of lab test values, where identical numerical results may reflect different health implications depending on demographic or physiological context. By including structured annotations of reference ranges along with contextual cues such as specimen type and patient attributes, LabQAR enables the evaluation of LLMs in tasks that require nuanced clinical reasoning. This makes the dataset particularly valuable not only for basic question answering but also for advanced applications like differential diagnosis, treatment monitoring, and personalized care.

Ultimately, LabQAR represents a step toward standardizing lab test interpretation using artificial intelligence. Traditional interpretation depends heavily on the clinician’s experience, which can vary widely. LabQAR provides a foundation for training and evaluating LLMs in a more consistent, explainable, and scalable manner, enabling improved clinical decision-making and enhanced patient outcomes. In addition to releasing LabQAR, we conducted experiments using state-of-the-art LLMs to identify lab test reference ranges. These experiments demonstrate the dataset’s utility and provide benchmark results to support future research and model development.

## Methods

The LabQAR dataset was created by compiling information from 550 laboratory tests gathered from authoritative medical resources, including Laposata’s Laboratory Medicine^37^, the American Board of Internal Medicine^38^, and Stanford Medicine^39^. Four annotators participated in labeling each lab test with crucial information, such as specimen type (e.g., blood, urine), gender or age variations, and other relevant factors like pregnancy or underlying conditions. Units were standardized to include both traditional and SI units, with reference intervals defined by lower and upper bounds. All laboratory tests were mapped to unique Logical Observation Identifiers Names and Codes (LOINC) identifiers using the LOINC clinical terminology system, accessible at https://loinc.org. This mapping provides standardized identification of laboratory tests within the dataset, enhancing consistency, enabling seamless integration with other healthcare datasets, and supporting accurate data exchange.

### Data Collection and Annotation

Figure 1 illustrates the data pipeline and evaluation framework for developing the LabQAR corpus focused on laboratory test reference ranges and interpretation. The process begins with the selection of 550 lab tests with reference ranges collected from authoritative sources, including Laposata’s Laboratory Medicine, the American Board of Internal Medicine, and Stanford Medicine.

**Fig 1.**
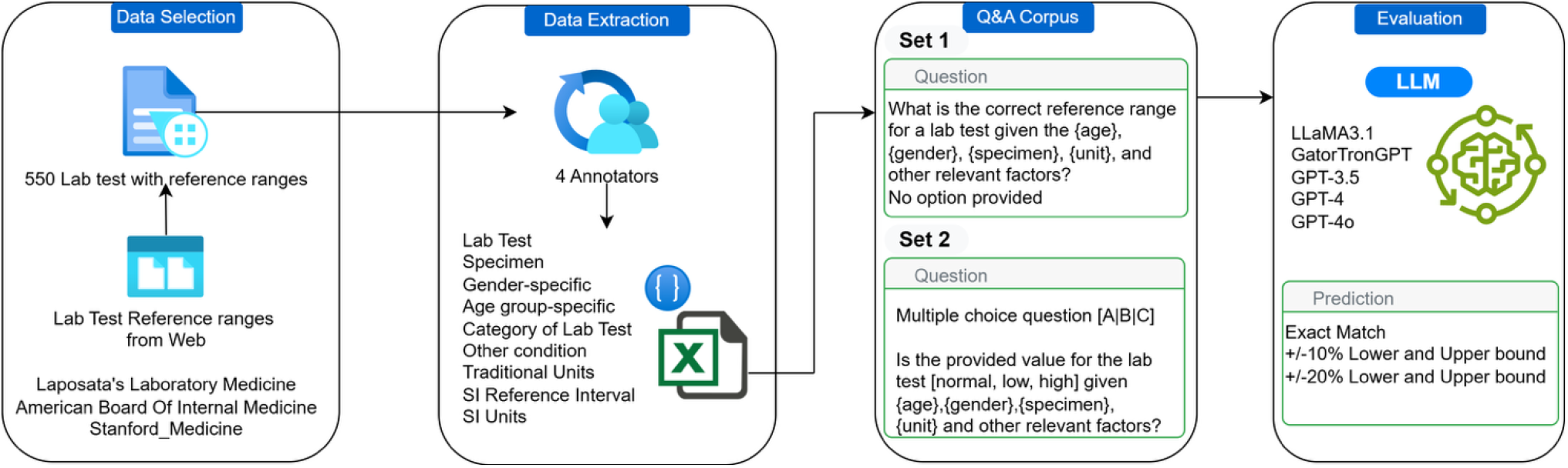
Overview of the LabQAR Dataset Construction and Evaluation Pipeline. The figure depicts the pipeline for creating and evaluating the LabQAR dataset. Reference ranges for 550 lab tests were curated from trusted medical sources and annotated by four experts for specimen type, age, gender, units, and clinical conditions. Two question sets were generated: Set 1 (open-ended) and Set 2 (multiple choice). These were used to evaluate LLMs (GPT-3.5, GPT-4, LLaMA3, and GatorTronGPT) based on exact matches and ±10% or ±20% deviation from reference values.

The dataset creation involved a multi-step process to ensure accuracy and consistency. First, the annotators underwent training to familiarize themselves with the data extraction guidelines (Supplementary Data 1) and the medical terminologies involved. This training phase was crucial to ensure uniformity in data annotation, minimizing inter-annotator variability. During the training, all the four annotators performed data extraction from the same set of lab tests from each of the three sources as their original formats differ. The PI met with the annotators once a week for five weeks to resolve conflicts. After training, each lab test entry was independently extracted by at least two annotators, followed by a consensus meeting to resolve any discrepancies. Four annotators manually extracted from each lab test detailed metadata such as lab test name, specimen type, gender-and age-specific values, test categories, conditions (e.g., menstrual phases), and measurement units (traditional and SI), along with their respective reference intervals. This rigorous approach ensured that the dataset maintained a high level of reliability and accuracy, which is essential for evaluating LLMs in a clinical context.

To curate and consolidate information for the same lab test from multiple sources, we systematically compared reference ranges, resolved discrepancies by prioritizing clinically validated and widely accepted values, and carefully documented any variations related to age, gender, or specimen type. Since Laposata’s Laboratory Medicine contains the most comprehensive set of lab tests, we assigned it the highest priority when selecting reference values. Laposata’s Laboratory Medicine lab test reference range compilation integrates values from multiple authoritative sources such as, Clinical Guide to Laboratory Tests (3rd edition Tests)^40^, SI Unit Conversion Guide^41^, American Medical Association Manual of Style (9th edition)^42^, Jacobs & DeMott Laboratory Test Handbook (5th edition)^43^, Clinical Diagnosis and Management by Laboratory Methods (20th edition)^44^, Kratz et al. in The New England Journal of Medicine^45^ and Tietz Textbook of Clinical Chemistry and Molecular Diagnostics (5th edition)^46^. This curated reference has been reviewed and updated by Jessica Franco-Colon, PhD, and Kay Brooks to reflect the latest clinical standards and consensus guidelines^37^. Based on this curated data and standardized extraction guidelines, annotators generated 550 detailed lab test reference ranges, covering various conditions, categories, and population-specific factors. These reference ranges encompassed 363 unique lab tests, ensuring broad clinical coverage. The structured dataset was subsequently used as input for a pipeline that generated the LabQAR question-answering dataset. The lab test reference range was mapped to a single, primary source to maintain consistency across the dataset. When multiple references are available for the same lab test, Laposata’s Laboratory Medicine was prioritized due to its comprehensive scope and clinical relevance. This allows us to anchor the data to a well-validated and widely used clinical reference. It is important to recognize that laboratory results can vary based on the testing method and institution. If any variations exist between multiple sources, our evaluation also accounts for these discrepancies by incorporating a margin-based assessment of model predictions. The models’ performance was evaluated based on exact matches as well as their ability to predict values within ±10% and ±20% of the reference bounds. This dual evaluation metric was chosen to reflect real-world clinical practice, where minor deviations from the reference range may still fall within acceptable diagnostic thresholds and not necessitate intervention.

### Creation of Q&A Sets about Reference Ranges of Lab Tests

Based on this structured data, two types of question-answering sets were developed: Set 1: open-ended questions asking for the correct reference range given patient-specific factors like age, gender, specimen, and unit; and Set 2: multiple-choice questions asking whether a provided lab value is ‘normal’, ‘low’, or ‘high’ based on those same contextual factors. The two sets of questions included in the dataset serve different but complementary purposes. For Set 2 questions, the goal is to assess a model’s ability to classify lab test results as ‘Low’, ‘Normal’, or ‘High’ based on contextual information such as specimen type, gender, age group, and clinical conditions. The Set 2 questions were generated by parsing the reference range for each lab test from curated data and generating a random test value within a realistic window (2×lower to 2×upper bound). This synthetic value is then classified using a deterministic logic: values below the lower bound are marked “Low,” those above the upper bound as “High,” and those within the range as “Normal.” Multiple-choice questions are automatically generated to include the clinical context and the randomly selected value, followed by shuffled answer choices (i.e., A, B, C). To ensure balance across answer options, the code tracks the distribution of correct answers and redistributes them evenly. This approach allows the creation of a robust, scalable evaluation set to test whether language models can interpret lab values accurately within nuanced clinical settings.

The open-ended questions are designed to test the LLMs’ ability to retrieve and synthesize information to provide precise reference ranges. These questions require the model to consider multiple factors, such as age and gender, making the task inherently complex. On the other hand, the multiple-choice questions aim to evaluate the models’ capacity for classification, which is critical in determining whether a given lab result falls within normal limits or indicates a potential health issue. Together, these question sets provide a comprehensive evaluation of the models’ abilities in both knowledge retrieval and clinical reasoning.

Figure 2 illustrates the process of transforming curated lab test metadata into structured question-answer pairs for a biomedical question-answering corpus. Following manual annotation by four trained annotators, key metadata—including lab test name, specimen type, gender specificity, age group, test category, traditional and SI units, and reference intervals—was compiled into a structured Excel sheet. A custom Python function was developed to convert this structured data into standardized JSON format for two distinct question types. Set 1 includes open-ended questions asking for the correct reference range based on specific contextual parameters, while Set 2 contains multiple-choice questions requiring classification of a provided lab test value as ‘normal’, ‘low’, or ‘high’. Two example questions demonstrate how features such as specimen type (e.g., plasma or serum), gender (e.g., male, any gender), age group (e.g., child, any age group), measurement unit (e.g., nmol/L), and reference ranges are systematically integrated into question formulation. This design enables consistent, machine-readable QA pairs that support downstream evaluation of language models on lab test interpretation tasks grounded in real-world clinical variation.

**Fig 2.**
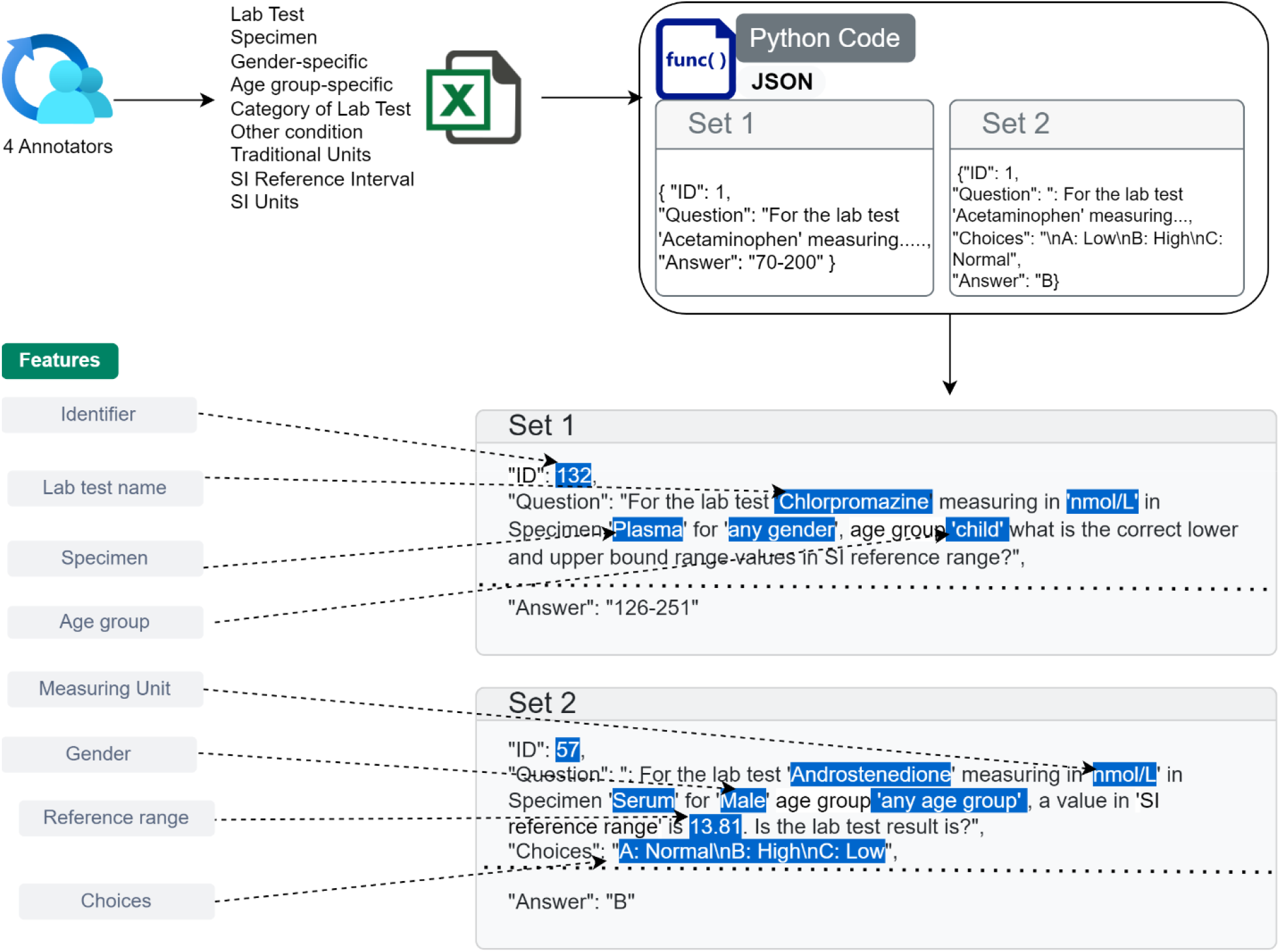
Generation of Structured QA Pairs from Annotated Lab Test Metadata. The workflow for converting manually annotated lab test information into structured question– answer (QA) pairs in JSON format. Four annotators extracted key metadata—including lab test names, specimen types, gender and age group specificity, reference intervals, and measurement units—which were compiled in an Excel spreadsheet. A Python function processed this structured data into two types of QA formats: Set 1 includes open-ended questions asking for the correct reference range based on contextual factors, and Set 2 includes multiple-choice questions asking whether a provided test value is ‘normal,’ ‘low,’ or ‘high.’ The highlighted examples illustrate how metadata fields such as test name (e.g., Chlorpromazine, Androstenedione), specimen (plasma, serum), age group, gender, and units (e.g., nmol/L) are used to formulate the question and determine the answer, supporting consistent and interpretable QA generation for model evaluation.

Additionally, the evaluation phase involved testing LLMs, including LLaMA 3.1^47^, GatorTronGPT^48^, GPT-3.5^16^, GPT-4^32^, and GPT-4o^49^, on their ability to predict both exact reference ranges and those within acceptable deviations (Set 1) and predict a given lab test value is as ‘Normal’, ‘Low’, or ‘High’ (Set 2).

## Dataset Characteristics

The LabQAR dataset comprises 550 lab test reference ranges derived from authoritative medical sources, encompassing 363 unique lab tests. Among these, 282 tests are general-purpose with no specific subgroup or condition, such as *Albumin*, which applies broadly across populations. A subset of 26 tests is tailored to gender-specific reference ranges, exemplified by *Apolipoprotein A*, with separate thresholds for males and females. Additionally, 7 tests exhibit age-specific variations, including *cholesterol* levels that differ between adults and children. The dataset also captures women-specific physiological contexts for 15 tests, such as *17*_α_*-Hydroxyprogesterone*, with stratification across the follicular, luteal, and postmenopausal phases. Furthermore, 33 tests are linked to other clinical conditions, like *Carboxyhemoglobin*, which is annotated with contextual values for nonsmokers and toxic exposure cases. This rich stratification enables precise, context-aware evaluation of large language models in clinical reasoning tasks. Table 1 summarizes the composition and subgroup characteristics of lab tests included in the LabQAR dataset, categorized based on patient-specific conditions such as gender, age, and physiological states.

**Table 1.**
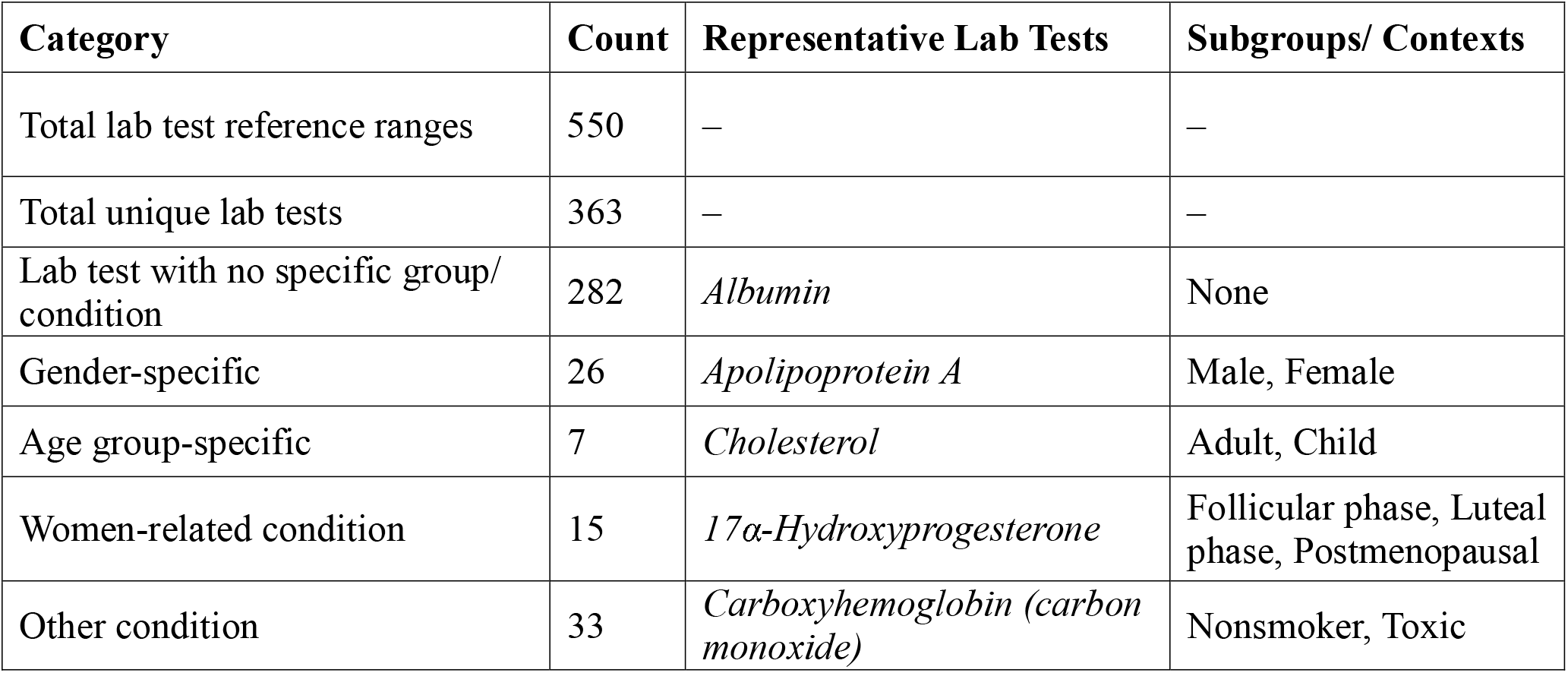
Summary of Lab Test Reference Range Categories in the LabQAR Dataset.

Figure 3 presents two sample question–answer (QA) pairs from the LabQAR dataset for the lab test *Follicle-stimulating hormone (FSH)* measured in IU/L in serum for postmenopausal females.

**Fig 3.**
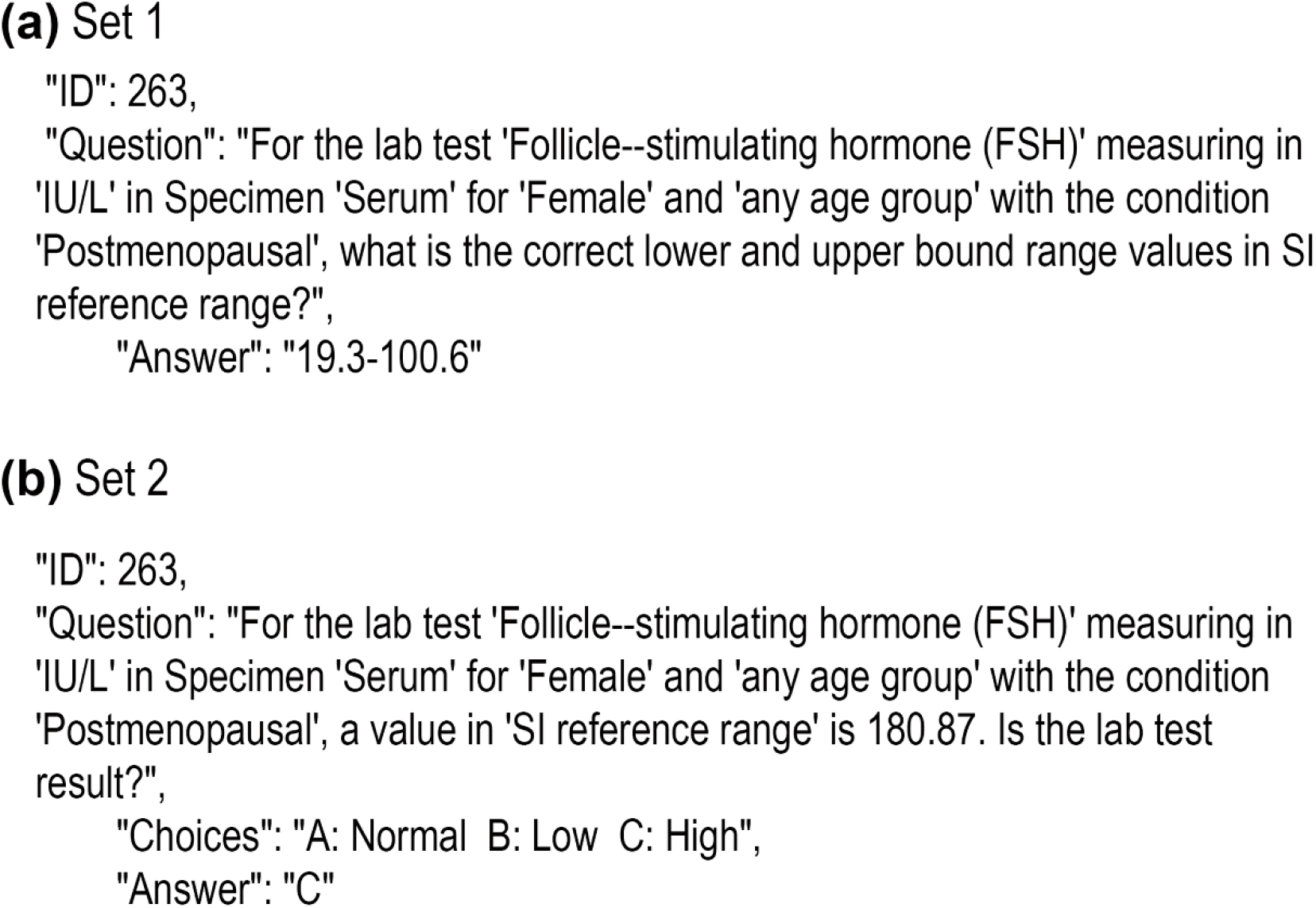
Examples of Q&A Pairs from LabQAR Dataset (a) Set 1 and (b) Set 2 for Follicle-stimulating hormone (FSH) Test under Postmenopausal Condition.

Figure 4 presents descriptive analyses of lab tests in the LabQAR dataset based on specimen types and reference range variability. Figure 4a shows the number of lab tests categorized by specimen type. *Serum* is the most common specimen (over 160 tests), followed by *whole blood, plasma*, and *serum/plasma* combined. Less frequently used specimens include *urine* and other specialized types. Figure 4b displays the relationship between the lower and upper bounds of SI reference ranges across lab tests. Most values cluster below 100 for both bounds, but a subset exhibits wide variability with upper bounds exceeding 1000, indicating the presence of highly variable lab measurements such as hormone or enzyme levels. This analysis supports downstream interpretation tasks by quantifying the diversity in specimen usage and numeric range distribution.

**Fig 4.**
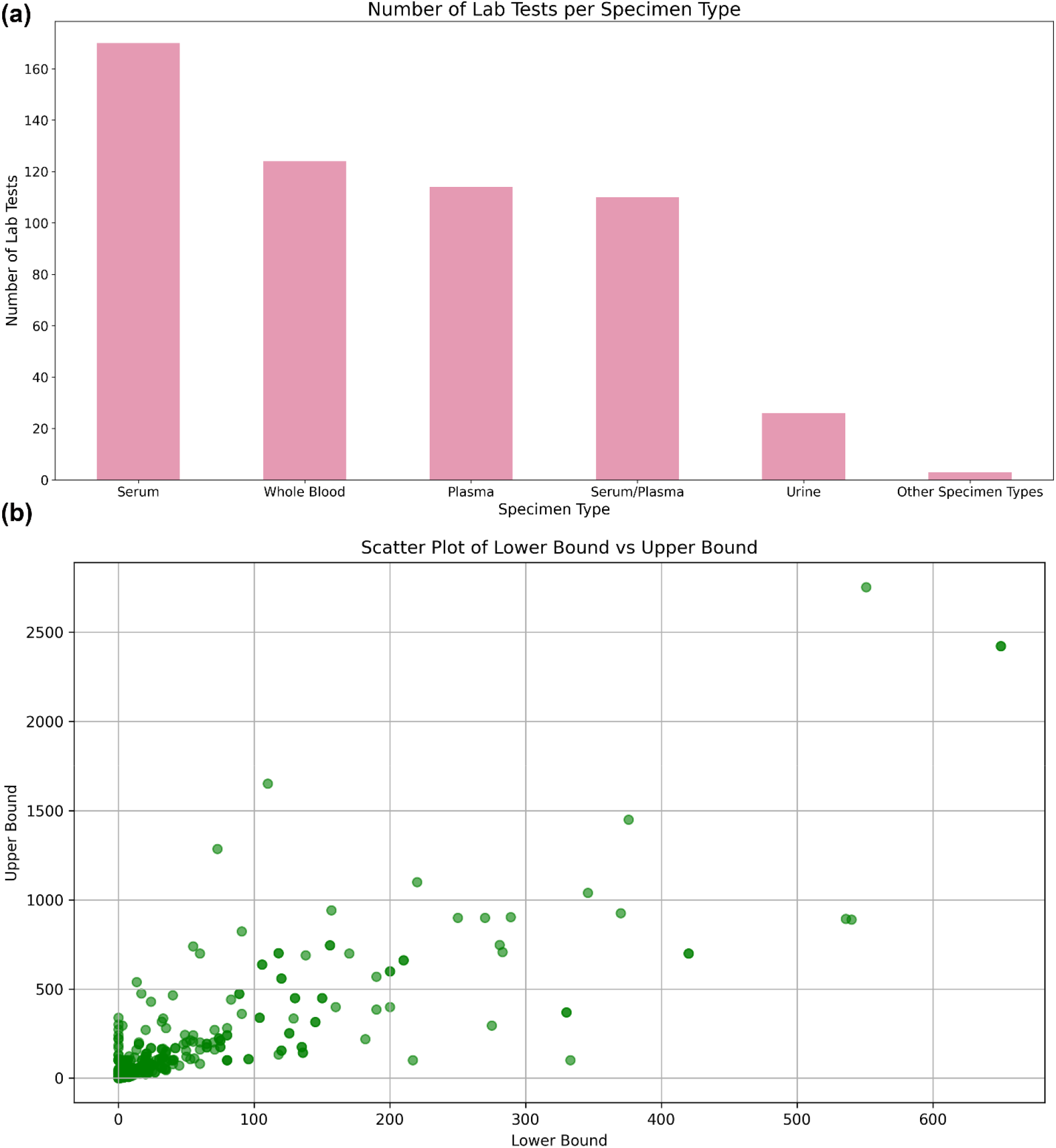
Distribution of Lab Tests by Specimen Type and Reference Range Variability Technical Validation.

In addition to releasing the LabQAR dataset, we conducted a series of technical evaluations using state-of-the-art LLMs to assess their ability to accurately identify lab test reference ranges and classify lab values based on contextual factors. Models evaluated include GPT-3.5, GPT-4, LLaMA3, and GatorTronGPT. These experiments provide benchmark results across two key tasks: (1) generating reference ranges given age, gender, specimen, and units, and (2) classifying provided lab values as ‘normal’, ‘low’, or ‘high’. Model outputs were evaluated using exact match accuracy and tolerance-based criteria of ±10% and ±20% around the gold-standard reference ranges. The results highlight the strengths and limitations of current LLMs in handling structured biomedical reasoning and demonstrate the utility of LabQAR as a standardized resource for training and evaluating future clinical NLP models.

## LABQAR Benchmarking

### Benchmarking metrics

To evaluate the performance of large language models (LLMs) on the LabQAR dataset, we employed a set of benchmarking metrics tailored to the two QA formats. For Set 1 (open-ended questions), model responses were assessed using exact match accuracy as well as tolerance-based measures: whether the predicted reference range fell within ±10% or ±20% of the ground truth lower and upper bounds. These metrics account for minor deviations while still emphasizing clinical precision. For Set 2 (multiple-choice classification), we used exact match accuracy to determine whether the model correctly classified the provided lab value as ‘normal’, ‘low’, or ‘high’ based on contextual attributes such as age, gender, specimen type, and units. Together, these benchmarking metrics provide a comprehensive view of LLM performance, capturing both numerical accuracy and clinical interpretability.

### Benchmarking results and discussion

We evaluated a range of large language models (LLMs) for our experiments, including LLaMA 3.1 (8B), GatorTronGPT, GPT-3.5 Turbo (gpt-3.5-turbo-0125), GPT-4 Turbo (gpt-4-0125-preview), and GPT-4o (gpt-4o-2024-08-06). The LLaMA 3.1 model was obtained from Hugging Face and deployed locally using the LangChain framework (langchain 0.3.25) on a system equipped with an NVIDIA RTX A6000 GPU with 48GB RAM. For local inference, the temperature was set to 0 to ensure deterministic outputs. The GPT-based models were accessed via the OpenAI API, with consistent parameter settings across all calls: temperature set to 0, top_p to 1.0, frequency_penalty and presence_penalty to 0.0, and a maximum token limit of 50 (adjusted as needed based on response length). These settings were selected to standardize model behavior and minimize randomness during evaluation. The results of the LLM evaluations are presented in Table 2. GPT-4o consistently outperformed the other models, achieving a 63.3% accuracy rate on multiple-choice classification tasks (Set 2), compared to GPT-4’s 43.1% and LLaMA 3.1’s 26.7%. Across all models, performance improved with relaxed criteria, particularly for the open-ended questions (Set 1). For instance, GPT-4 turbo improved from 27.8% exact matches to 49.8% accuracy within 20% bounds.

**Table 2.**
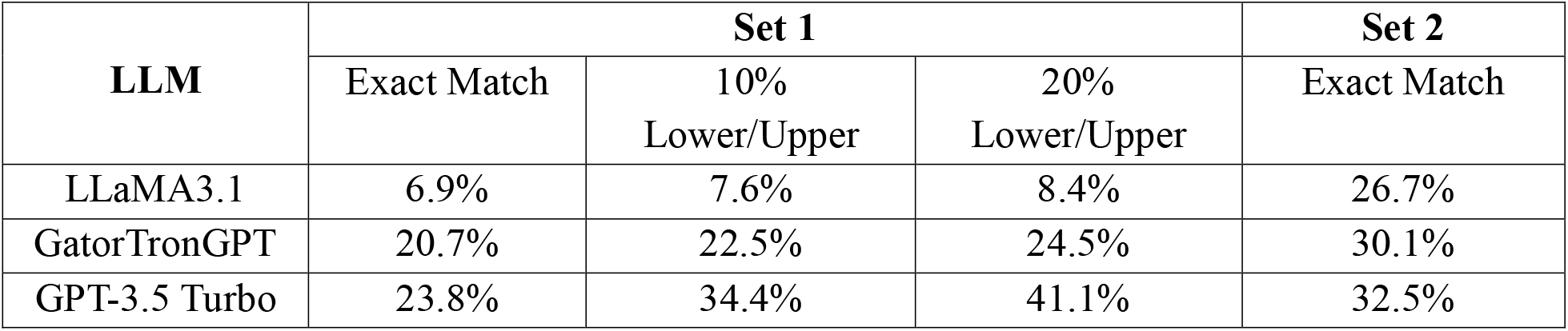
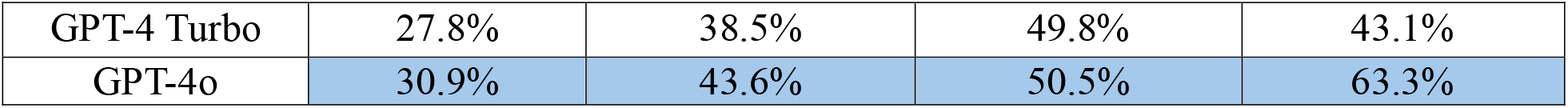
Benchmarking Metrics for Evaluating LLM Performance on LabQAR Question Sets.

The performance differences observed between the models can be attributed to several factors. GPT-4o’s architecture and training data were optimized to better handle domain-specific information, which likely contributed to its superior performance. In contrast, LLaMA 3.1’s relatively lower accuracy indicates challenges in capturing the nuances of lab test reference ranges, particularly when contextual factors like age and gender are involved. This highlights the importance of specialized training and domain-specific datasets for improving model accuracy in healthcare applications.

Interestingly, the results also showed that relaxing the criteria for accuracy significantly improved the models’ performance across the board. This suggests that while exact match prediction remains challenging, LLMs are capable of providing clinically useful approximations. For clinical practice, this means that while models like GPT-4o may not always provide perfect answers, they can still offer valuable insights that could assist clinicians in making informed decisions. Such findings underscore the potential utility of LLMs as tools to support, rather than replace, human expertise in healthcare.

The evaluation revealed that GPT-4o accurately predicted reference ranges for tests such as Chloride and Hydroxyproline, while LLaMA 3.1 struggled with tests like Glucose and Follicle-Stimulating Hormone (FSH). GPT-3.5 and GPT-4 performed well in predicting common tests, including Glucose, Hemoglobin A1c, and Albumin. Notably, GPT-4o’s higher accuracy indicates its ability to generalize across various laboratory tests, making it a promising candidate for clinical applications.

One of the main challenges highlighted by the evaluation was the ability of LLMs to manage overlapping or age-specific reference ranges. For example, GPT-4o demonstrated notable accuracy in predicting reference ranges for Chloride, which has relatively stable limits across age groups. However, for more complex tests like FSH, which vary significantly depending on age and gender, even GPT-4o faced difficulties. This points to the need for further optimization of these models, potentially through the integration of more specialized medical datasets or enhancements in contextual understanding mechanisms.

Another significant finding from the evaluation was the impact of training data quality on model performance. The superior performance of GPT-4o can likely be attributed to the diversity and quality of its training data, which included more domain-specific medical texts. This indicates that future advancements in LLM capabilities could be achieved by expanding training datasets to include a wider variety of clinical data, such as electronic health records and detailed medical guidelines. Additionally, fine-tuning models on specific tasks, such as reference range prediction, could further enhance their applicability in clinical environments.

Despite its strong performance, GPT-4o’s accuracy of 63.3% indicates that significant improvements are still necessary for these models to be effectively integrated into clinical workflows. Future enhancements, such as the integration of retrieval-augmented generation (RAG) using public data sources like MedlinePlus, could further improve accuracy and reliability. Incorporating real-time data retrieval could allow models to access the most current medical knowledge, ensuring that their predictions remain relevant as medical guidelines and standards evolve.

## Conclusion

LabQAR provides a robust dataset that supports accurate predictions and classifications of laboratory test results, offering valuable insights for clinical decision-making. This dataset is a significant step toward improving automated interpretation of lab tests and enhancing the capabilities of LLMs in personalized healthcare. By providing detailed annotations and reference ranges, LabQAR enables LLMs to better understand complex medical data, ultimately supporting clinicians in delivering more informed and timely patient care.

## Usage Notes

The accompanying README file provides detailed instructions for processing the LabQAR dataset. This dataset is specifically designed to support a range of clinical NLP tasks, including: Reference Range Extraction – retrieving the correct lower and upper bounds for laboratory test values based on test name, specimen type, measurement unit, and demographic context. Lab Result Classification – interpreting a given lab test result as High, Normal, or Low by comparing it against the appropriate reference range. These tasks enable the development and evaluation of models for clinically informed question answering and result interpretation in laboratory diagnostics.

## Data Availability

All data produced in the present study are available upon reasonable request to the authors.

## Code Availability

The code used to prepare the LabQAR dataset is provided at https://github.com/balubhasuran/LabQAR, and the source code for the benchmarked experiments can be found with the dataset.

## Acknowledgments

This work was supported by the Agency for Healthcare Research and Quality grant R21HS029969 (PI: ZH). This project was also partially supported by the University of Florida-Florida State University Clinical and Translational Science Award, which is supported in part by the National Center for Advancing Translational Sciences under award UL1TR001427. QJ and ZL were supported by the NIH Intramural Research Program, National Library of Medicine. We would like to thank the four undergraduate students Angelique Deville, Hailey Thompson, Maggie Awad, and Yash Alva for their work in extracting from lab test documents.

## Competing Interests

The authors declare no competing interests.

## Notes

### Competing Interest Statement

The authors have declared no competing interest.

### Author Declarations

The source data is publicly available.

