## Supplementary Data 1 for "LabQAR: A Manually Curated Dataset for Question Answering on Laboratory Test Reference Ranges and Interpretation"

**Lab Test Details Extraction Guidelines**

**Objective**

To extract standardized metadata from lab test documents and encode them into a structured Excel format for two types of clinical question-answering (QA) datasets:

- Set 1: Reference range retrieval (open-ended)
- Set 2: Lab result classification (multiple-choice)

**1. General Instructions**

• Review each row in the source table (e.g., PDF or webpage).

• Annotate all relevant information per lab test, including units, reference ranges, and contextual conditions.

• Maintain consistency with terminology and formatting for downstream automation (e.g., JSON conversion).

• Use drop-down options or controlled vocabularies wherever available (e.g., 'Serum', 'Plasma', 'Male', 'Female', 'Child', 'Adult').

**2. Required Metadata Fields to Annotate**

| Feature | Description | Example Value |
|---|---|---|
| Lab Test Name | Full name of the lab test | Acetaminophen (therapeutic) |
| Specimen Type | Biological sample used | Serum, Plasma |
| Gender | If reference ranges are gender-specific | Male, Female, Any |
| Age Group | If reference ranges vary by age | Adult, Child, Any |
| Measurement Units | Both Traditional and SI units | μg/mL, μmol/L |
| Reference Range | Traditional and SI reference intervals | 70–200, <0.1 |
| Conversion Factor | Factor between traditional and SI units | 6.62 |
| Test Conditions | Specific physiological or clinical conditions | Luteal phase, Fasting |
| Category | Type of lab test | Therapeutic Drug Monitoring |
| Source Reference | Source of extracted data | Laposata, ABIM |

### 3. Special Handling Cases

• Multiple Specimens: Separate entries or note both specimens using a delimiter (e.g., 'Serum, Plasma')

• Range with '<' or '>' signs: Keep original format (e.g., '<0.1') and annotate carefully

• No SI Units: Leave blank or indicate 'N/A'

• Ambiguous Units or Ranges: Flag for discussion or verification

### 4. Generating QA Pairs

**Set 1: Open-ended question (Reference Range Retrieval)**

Template: For the lab test {Lab Test}, measuring in {SI Unit} in {Specimen} for {Gender}, age group {Age Group}, what is the correct lower and upper bound range values in SI reference range?

Answer: {SI Reference Range}

**Set 2: Multiple-choice question (Lab Result Classification)**

Template: For the lab test {Lab Test}, measuring in {SI Unit} in {Specimen} for {Gender}, age group {Age Group}, a value in SI reference range is {Value}. Is the lab test result normal, low, or high?

Choices: A. Normal, B. High, C. Low

Answer: (e.g., B)

### 5. Critical Annotation Examples

| Lab Test | Specimen | Gender-specific | Age grou specific | Women-related condition | Types of reference range | Traditional Reference Interval | Tradition Units | Conversio Factor, | SI Reference Interval | SI Units |
|---|---|---|---|---|---|---|---|---|---|---|
| 17α- -Hydroxyproge erone | Serum | Female | - | Follicular phase | Normal | 15–70 | ng/dL | 0.03 | 0.4–2.1 | nmol/L |
| Follicle-- stimulating hormone (FSH | Serum | Female | - | Ovulatory phase | Normal | 6.17–17.2 | mIU/mL | 1 | 6.1–17.2 | IU/L |
| Alanine | Plasma | - | Adult | - | Normal | 1.87–5.88 | mg/dL | 112.2 | 210–661 | μmol/d |
